# Public attitudes towards COVID-19 vaccination in children: A qualitative study

**DOI:** 10.1101/2021.07.28.21261252

**Authors:** Simon N Williams

## Abstract

**Background:** COVID-19 vaccinations in children remains controversial. In the UK, as of August 2021, they have not been approved, except in a few limited circumstances. To date, little qualitative research exists to explain the reasons and nuances behind public attitudes on this issue.

**Methods:** Qualitative group and one-to-one online interviews were conducted with a diverse sample of 24 adults to explore their views on the issue of COVID-19 vaccination in children.

**Results:** COVID-19 vaccination in children was framed as a complex problem (a “minefield”). Six themes emerged to explain participants views: (1) Uncertainty over whether children can catch, transmit or be severely harmed by COVID-19; (2) Lower risk tolerance for unknown longer-term effects of the vaccine in children; (3) Association of the vaccine program with government’s handling of the pandemic; (4) Local social norms as a driver of hesitancy; (5) Vaccinating children as a way to protect vulnerable adults; (6) Children’s vaccination as parental choice.

**Conclusions:** Public attitudes to COVID-19 vaccination in children are likely to be met with more hesitancy compared to adult vaccinations. Public health communications will need to combat this hesitancy if vaccine uptake for children is to be pursued as a public health policy.

## BACKGROUND

Approximately one-and-a half years into the novel coronavirus (COVID-19) pandemic, the virus continues to exert a considerable toll on global mortality and morbidity, although the development and roll out of vaccines offer much hope for the future.^1^. In the UK, as of July 25^th^ 2021, a total of 83 million COVID-19 vaccinations had been administered.^2^ However, despite the relative success of the adult vaccination programme, questions over how, and indeed whether, ‘herd immunity’ can be achieved for COVID-19, particularly in light of new more transmissible variants (e.g. the Delta variant) remain, including whether or not children should be vaccinated towards this end.^3^

There is little academic consensus as to whether or not children should be vaccinated, in part due to the novelty of the disease and the vaccines, and in part due to wider ethical, practical and social considerations. As well as the potential contribution to herd immunity, arguments in favour of vaccinating children against COVID-19 include: preventing rare but severe disease in children; reducing transmission from children to adults; priming children’s immune response to future (re-)infection; and helping to keep schools open.^4^. Arguments against, tend to focus on the fact that children are significantly less prone to serious outcomes from COVID-19 and that it is necessary to obtain substantial safety data before widespread use amongst (non-clinically-vulnerable) children.^5^

Although many opinion pieces include or focus on the more ethically-challenging issue of mandatory vaccination in children – something acknowledged as being unlikely or even counter-productive in the short-term ^4 5 6^ However, many of the arguments raised (for and against mandatory vaccination) are also relevant for voluntary vaccination in children. A number of criteria for determining whether or not children should be vaccinated against COVID-19 have been proposed, including: disease related criteria, including the extent to which it would reduce mortality, morbidity, and transmission in the population); vaccine-related criteria, including what they take to be the most important criterion – evidence pertaining to the safety of the vaccine in children; and implementation-related criteria, including is acceptable to the medical community and the public. ^7^ This latter criterion is the focus of this article – namely to explore the perspectives of members of the UK public (including but not limited to parents) on the issue of COVID-19 vaccination in children.

A growing number of public opinion surveys are being conducted, which find that, although public attitudes to the issue are mixed, a majority of the public overall, and parents specifically, are in favour of voluntary vaccination of children against COVID-19.^8 9 10^. Surveys are beginning to document the reasons behind these views. Reasons in favour of vaccinating children included: to prevent the spread of COVID-19; to prevent their children from catching COVID-19; to get back to normal more quickly. Reasons for opposition or hesitancy included: concerns over long-term effects of the vaccine on their child’s health, concerns over side-effects, a lack of knowledge of whether the vaccines had been tested in children, children are too young; and children are unlikely to get very ill from COVID-19.^10^

Qualitative evidence can provide context, nuance and depth to emergent findings on public attitudes. This paper explores the participants attitudes towards COVID-19 vaccination in children, including the reasons behind their views.

## METHODS

Participants were recruited as part of the qualitative component of an ongoing, longitudinal mixed methods study exploring public views on the COVID-19 pandemic in the UK. More details about the methodology can be found in previous publications.^11 12 13^. In this paper, we report on data from a rapid round of 4 focus groups and 3 one-to-one interviews with a total of 24 participants between 1^st^ July – 25^th^ July 2021. Participants were initially recruited to the study from March-July 2020 and were all UK-based adults aged 18 years or older. Recruitment for the study took place via a combination of social media advertising and snowball recruitment. Purposive sampling was used to seek as diverse a range of ages, genders, race/ethnicities, UK locations, and social backgrounds as possible, although the limitations of the final sample are discussed below as well as in previous publications.^11 12^ Full demographic summary details are provided in Table 1.

**Table 1:**
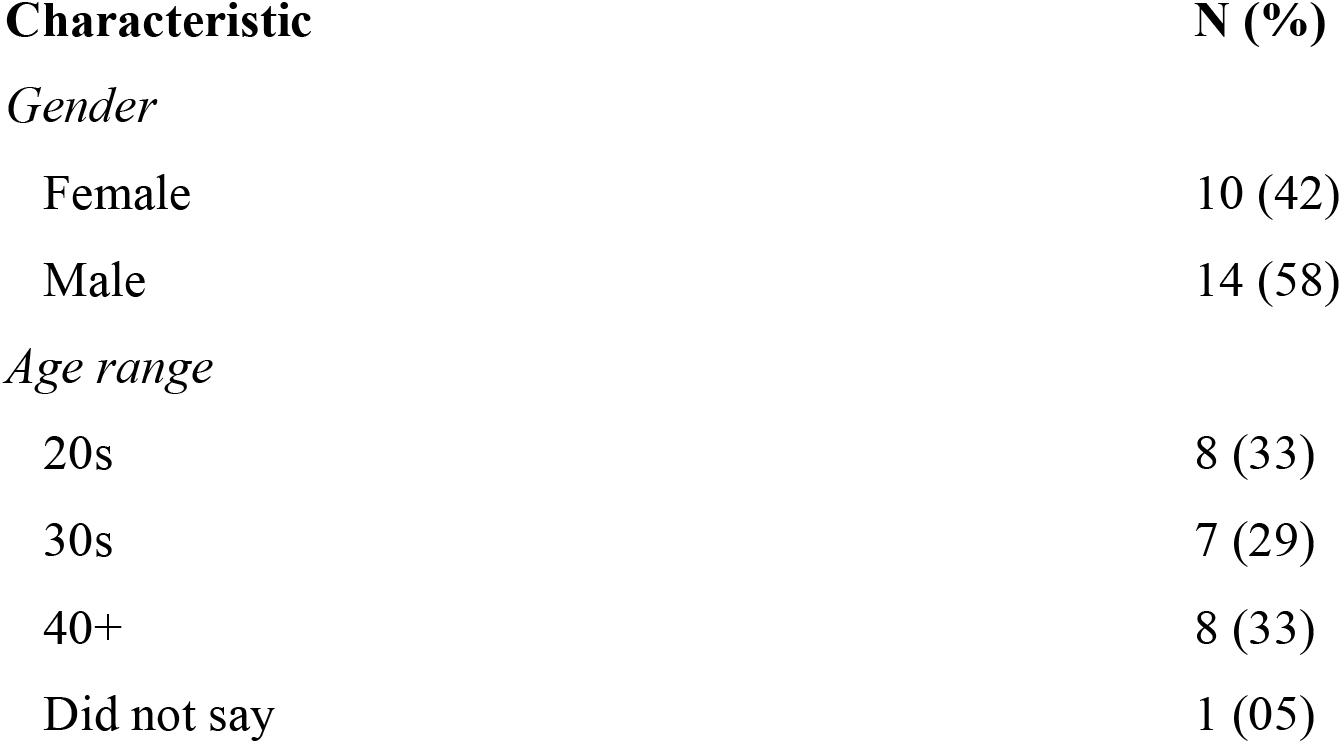
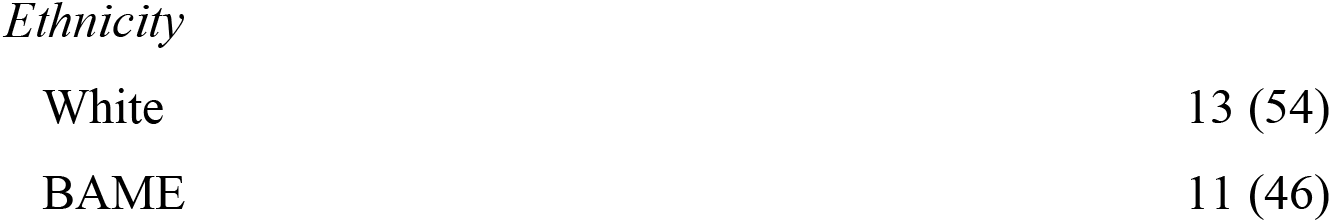
Demographic characteristics for participants in this report

All interviews took place remotely via videoconferencing (Zoom) and lasted approximately one hour. Questions were guided by a semi-structured schedule built around the research question and literature above (particularly focused on participants’ reasons for their views on whether or not they were favourable towards COVID-19 vaccination in children). Ethical approval for the study was granted by Swansea University’s School of Management and College for Human and Health Sciences research ethics committees and all participants gave informed consent and had their data anonymized.

Data were analysed in accordance with a framework analysis approach.^14^. Data were coded and organized according to themes and subthemes, using a data matrix (framework) that was developed primarily inductively during the early interviews and then applied and modified during the remaining interviews.^14^ Coding was performed using NVivo (version 11.4.3, QRS).

## RESULTS

### COVID-19 vaccination in children as a “minefield”

Overall, the issue of COVID-19 vaccination in children was framed as a complex issue with no clear-cut answer to the question of whether they should be offered. Although there was a spectrum of views represented, with some participants being strongly opposed to the idea and others being largely in favour, most seemed to frame it as a “tricky” and “a grey area”. Although the specific considerations that participants were weighing up will be discussed separately below, it is important to point out that participants often integrated them into one answer when trying to account for their views – quite literally articulating (‘thinking out loud’) the complexity of the issue as they saw it, and the fact there was to them no clear-cut answer. The following quote illustrate the full extent of the complexity and uncertainty that many participants also exhibited, as they wrestled with the question over whether COVID-19 vaccines should be offered to children:

> “I just think it’s a grey area. I can’t really decide which way is best to be honest, there’s like pros and cons to each side. … People say like you’ve got to vaccinate kids because they’re obviously going to grow and then they’ll be part of society unvaccinated. But then you are subjecting kids to unnecessary injections potentially. And about consent as well, you know you are kind of forcing the injection on the kid. And considering how hazy the UK government has been with their decisions, it is another one that’s hard to decide, I think they will flip-flop again. There is no easy answer. … They’ll have to research, maybe get parents opinions on it. I guess indirectly, if they’re not vaccinated, it can affect everyone because kids could potentially spread it, you know between themselves, but then again should we label kids as these germ spreaders. … I don’t really think it’s fair because you know adults spread it just as much. I guess what I’m trying to say is, I’m unsure on the whole matter. I think it’s a minefield.” (Participant 1, Male, 30s)

The above quote, which was taken from one response reveals the participant’s complex stream-of-thought as he struggles to decide what the ‘right’ answer is. It captures many of the factors (and themes) influencing participants indecisiveness, uncertainty or hesitancy around whether or not COVID-19 vaccines was a good thing, (discussed in more detail in the following sections), including: the idea that children are not at particularly high risk from serious illness and uncertainty around their role in transmitting the virus (“you are subjecting kids to unnecessary injections potentially”) (see: *Uncertainty over whether children can catch, transmit or be severely harmed by COVID-19*); the problem of agency, consent and responsibility, specifically whether young children should be “forced” to have vaccines (“consent as well, you know you are kind of forcing the injection on the kid” and “They’ll have to research, maybe get parents opinions on it”) (see: *Children’s vaccination as parental choice*); connecting a general lack of trust in government to a lack of confidence in the (safety and use) of COVID-19 vaccines lack of trust in government (“considering how hazy the government has been with their decisions”) (see: *Distrust in government/trust in science*); the idea of childhood vaccines as a way to contribute to overall population (‘herd’)immunity (“you’ve got to vaccinate kids because they’re obviously going to grow and then they’ll be part of society” and “if they’re not vaccinated, it can affect everyone because kids could potentially spread it”) (see: *vaccinating children as a way to protect society*).

For parents, the issue was seen to be equally if not more complex, as they wrestled with the as-yet unresolved dilemma of whether or not to vaccinate their *own* child – a decision which had a strong affective dimension to it (“I do feel quite mixed”), and which was seen to be a “big responsibility”:

> “I do feel quite mixed about it … the disruption that the pandemic has had on a child’s education, you know whole year groups having to isolate because one child tested positive, I think that can’t continue and it’s just having a huge impact on their education as well. If there was enough evidence that they [vaccines] were safe and the risk of the vaccine outweighed them not being vaccinated then, yes, I would [vaccinate my children]. I think it would depend on what the research is saying, it’s still early days and you feel like it’s a big responsibility making those decisions for your children and their health. The other vaccines … have been around for a long time the childhood immunizations, tried and tested, and you do need to start somewhere but it feels like is it necessary for them, when it’s not initially affecting children. But then you have got this delta variant which does seem to be have more children testing positive. … It’s hard because you feel like you need a bit of time to see, but then in saying that it’s like your expecting other people’s children to be vaccinated so that you’ve got the data. … It’s important to do what we can to get out of the pandemic situation but it much harder when it’s *your* children. … I have very mixed feelings about it.” (Participant 15, Female, 30s)

Again, many of the main factors (our themes below) were seen to interconnect as participants sought to weigh up the risks and benefits, in light of what was seen to be considerable uncertainty, including: the extent to which children ‘need’ the COVID-19 per se (“is it necessary for them, when it’s not initially affecting children?” and “but then you have got this Delta variant which does seem to have more children testing positive) (see: *Uncertainty over whether children can catch, transmit or be severely harmed by COVID-19);* the extent to which children should be spared or protected more from any potential, unknown, long-term side effects from the vaccines (“other vaccines … have been around for a long time the childhood immunizations, tried and tested” and “it’s still early days”) (see: *Lower risk tolerance for unknown longer-term effects of the vaccine*); the idea that vaccinating children might help bring an end to the pandemic (“It’s important to do what we can to get out of the pandemic situation”) (see: *Vaccinating children as a way to protect society);* and the burden of responsibility - of having to decide something that affects your own child or children as well as others (“It’s hard because you feel like you need a bit of time to see, but then in saying that it’s like your expecting other people’s children to be vaccinated so that you’ve got the data”) (see: *Children’s vaccination as parental choice*). The latter will be discussed in terms of individual versus collective responsibility (including the potential role of NIMBY syndrome (see: *Discussion*)). These factors will be discussed in more depth in the following sections.

The above quote also illustrates a consideration that was raised by some parents and those working with young children (but which was not frequent enough to be considered a main theme) which was whether any potential, unknown, long-term harms of the vaccine in children would be outweighed by the short-term harms caused by the disruption to children’s lives due to enforced school closures and absenteeism as a result of isolation policies (“it’s just having a huge impact on their education as well”).

Indeed, most participants saw the issue as one of weighing up the relative benefits and risks, often struggling to come to a conclusion over what the “right” answer was. One of the main reasons as to why it was seen as such, is because it was acknowledged by many as ethically challenging– for example being seen as a “sensitive topic”:

> “I think, for me, as a parent, that is a very, very sensitive topic is going to because, even for me as an adult if I find it scary to actually go for this jabs, I’d be very reluctant to take a decision to go for vaccination in children.” (Participant 9, Male, 40s)

Interestingly, very few participants expressed *unequivocal* or unqualified support for vaccinating children. This contrasts with our earlier research on adult vaccinations, where a number of participants were unequivocally in favour of COVID-19 vaccines and vaccines in general (categorized as ‘full accepters’).^15^ Some however, did express largely positive views, although these were more likely to be non-parents who couched or qualified their support in two ways: Firstly, that they had less stake in the issue (because they themselves didn’t have children) and, secondly, their support was conditional on the vaccine having been proven to be safe and efficacious specifically in and for children. These qualifications are discussed in more detail below.

All seven parents in our study expressed hesitancy and concerns, with one stating outright they did not agree with vaccinating children against COVID-19 (“No definitely not, one hundred percent no” (Participant 11, Female, 30s). Perhaps unsurprisingly, those who themselves had refused the vaccine and held generally negative views about adult COVID-19 vaccines in adults (identified as ‘refusers’ in our previous study,^15^ were clearly and strongly opposed to the very idea of COVID-19 vaccination for children (“Me personally, I don’t support vaccinating children against Covid” (Participant 9, Male, 20s)). However, some participants who were quite favourable towards COVID-19 vaccination in adults (and who themselves had been vaccinated) were quite strongly opposed to the idea of vaccinating children:

> “I find it quite sinister. … As much as I have been for the vaccine, the rhetoric has switched and changed since its inception, where firstly it was something for the elderly, and then the middle-aged and its encroached on different age groups almost by stealth, and now we are talking about vaccinating our children. … I think it’s too much, it’s not necessary and it opens Pandora’s Box. It goes into a different sphere altogether when we start to promote the obligatory vaccination for our kids on top of all the other things they get jabbed for (Participant 13, Male, 40s)

As the above quote suggests, some felt that vaccinating children was going “too far” and was “not necessary”. The reasons behind participants opposition to or hesitancy concerning COVID-19 vaccines in children will be discussed in more detail in the following section. Interestingly, the above quote also illustrates the fact that some participants tended to conflate the question of whether or not the COVID-19 vaccine should be made *available* (i.e. optional) to children with the question of whether or not children should *be* vaccinated per se. Participants were not asked whether mandatory vaccination should be introduced (since this is not a realistic policy scenario in the UK). Nevertheless, some participants responded to the question by arguing that “obligatory” vaccination in children was not ethical or desirable. This is a subtle but important distinction since there may be the misassumption in some people’s minds that approving the vaccine’s use in children equates to mandating its use in children. As discussed below, communicating and emphasising the fact that childhood vaccinations, if approved, will be optional and not mandatory might help improve their overall acceptability amongst those who might mistakenly assume they are being considered for mandatory use.

### Uncertainty over whether children can catch, transmit or be severely harmed by COVID-19

One prominent theme concerned participants’ uncertainty over the extent to which children could either themselves catch, suffer from, and transmit COVID-19. To an extent the public’s uncertainty over these issues was seen to be (and in some senses was actually) a reflection of scientific uncertainty over the same issues (although the public’s perception of scientific uncertainty was at times magnified or distorted). For example, some participants felt that they didn’t know whether children played a significant role in transmitting COVID-19. This led to some to argue that, because of this uncertainty, they weren’t sure if vaccinations were necessary or a good idea. Some suggested that there had been conflicting or confusing evidence or data (including that which they had read or heard about via the media and via their own personal or anecdotal evidence). This included those working in front line roles working with children, for example in education and social care roles:

> “It’s a tricky one. I think there’s so much like discrepancy on that the data with covert in children, I still personally don’t know whether they are, I mean there was all that all that talk about six months ago that they were like super spreaders and actually children in schools were spreading it. And then all of a sudden, it was that they weren’t super spreaders and actually they were fine and it was the adults and then you know all of this kind of back and forth, so I still don’t even know like when I’m teaching if kids are spreading the virus. If children were, you know, spreading the virus that in such close contact with each other, throughout the day. Whenever we seem to lock down, you know singular classes it’s always like one case, it’s never like large numbers that have got Covid.” (Participant 3, Male, 20s).

> “I am getting less and less children coming into school. I felt like most of the tests are coming back negative and so they haven’t got Covid. So that makes me question do they really need the vaccine if they haven’t got it.” (Participant 5, Female, 20s)

Ultimately this could be seen as a matter of risk perception in relation to the perceived benefits. That is, whereas many participants were aware of the benefits of vaccinating adults (particularly older and vulnerable adults),^15^ participants were less clear over or confident in the benefits of vaccinating children.

Those who were particularly opposed to (refusers) vaccination in children, were more likely to unequivocally argue that COVID-19 was something that children were at risk of dying from or being “severely impacted” by, or was even something they were particularly “prone” to (i.e. they did not perceive it as a complex or scientifically uncertain issue):

> “I just don’t *feel* kids need the vaccine, they not had the virus anyway, they are just not prone to it” (Participant 11, Female, 30s).

> “I don’t support vaccinating children against common because I don’t see the risk of them dying or being severely impacted biologically is that great so no I don’t support it at all.” (Participant 10, Male, 20s)

Some other participants (who were hesitant but not necessarily opposed) also made the related argument that children didn’t need the vaccine because they had “young” and healthy immune systems and were more able to fight the virus “naturally”. This was also seen to be a key factor in accounting for hesitancy in regard to adult vaccinations.^15^ As one participant put it, not putting the vaccine “in the body” was part of keeping healthy, which should be “sufficient” to protect against the virus:

> “So I have a kind of *feeling* that for youngsters that the when you are young natural immune system is really strong … and that if you take care of your lifestyle and eat healthy that should, for now be sufficient than actually going for this job if you don’t know what it can do to you in the body. (Participant 9, Male, 40s)

A number of the above quotes suggest that participants’ views were largely affective in nature (i.e. the affect heuristic)^16^ – that is as a result of a general “feeling” they had. As will be described below, this short-term affective judgment may have been tied to their belief that there was not enough information at present for them to make a more informed (‘rational’) choice on the issue of COVID-19 vaccinations in children.

Some participants distinguished between younger and older children. They were inclined to be more favourable to the idea of vaccinating older children (i.e. teenagers) for two reasons: firstly, because as they understood it, teenagers were more likely to transmit the disease compared to younger children (“they are out and about a bit more” (Participant 4, Male, 40s)), and secondly, because as they understood it younger children were more at risk of any potential longer-term side effects (a theme that will be explored in more detail below):

> “I think I would be much more confident saying like teenagers, 15, 16, even 13 to 18, I think there is evidence that they spread the virus. And because they are older it is more accepted. With younger children, everyone is like, ‘think of the children, we don’t know the long-term effects’. With older teenagers there is definitely a differentiation … it makes more sense in a secondary [school] context” (Participant 3, Male, 20s).

### Lower risk tolerance for unknown longer-term effects of the vaccine in children

At the same time as participants were uncertain or lacked confidence in the benefits of COVID-19 vaccines in children (at least relative to the perceived benefits for vaccinating adults), a number also felt that the risks may be higher in children. This included some of the parents in the group, who exhibited a lower risk tolerance for vaccinating their children. Parents tended to frame their decision in terms of a risk-benefit trade-off, with some tending to feel that the perceived risks of the vaccine outweighed the perceived benefits:

> “I feel quite apprehensive, mine [her children] are [aged] 8 and 11. Locally, I’ve not really known any children who’ve been seriously ill with it, and that it feels that possibly for this age group the risks of the vaccine are possibly higher than the risks of them if they were to have Covid” (Participant 8, Female, 30s)

> “Although I have been vaccinated, I wouldn’t want my son to be vaccinated. Although there has been research done, I know it is quite early days., so I would rather take the risk of him getting Covid than the risk of him having the vaccine … Although people think it’s safe, I still feel that some point in the future they will discover something [about the vaccine] that affects children more than adults (Participant 15, Female, 40+)

These perceived risks invariably centred around the concern that there might be unknown, longer-term side effects related to the vaccine. This echoes findings from our previous study on attitudes towards adult vaccinations, where concerns over long-term side effects were a key driver of hesitancy.^15^ As far as views on children were concerned, there was a perception that they were perhaps *more* susceptible to potential longer-term side effects by virtue of their age and the fact that they were still developing. For some, the lower risk tolerance, and this perception that children were more “vulnerable” to potential side effects, led them to be explicitly opposed to the idea of a vaccine in children (“I don’t think there is a need for any type of fluid going into a child’s body”):

> “There needs to be another initiative [apart from COVID-19 vaccinations in children] to improve [sic] a reduction in [COVID-19 case] rates. I don’t think there is a need for any type of fluid going into a child’s body. Even with the parent’s consent, because children’s health could be severely affected. Because even with adults, the side effects you’ve noticed from taking the vaccine and children are more vulnerable and more [at] risk so don’t I agree with the idea” (Participant 2, Male, 30s)

The implication here is that some people may believe (perhaps somewhat contradictorily) that while children are *less* likely to be severely impacted by COVID-19 because of their “biology” (i.e. because they have “strong immune systems”) they are also *more* likely to be severely impacted by COVID-19 vaccines because of their biology (i.e. because their bodies are still developing). Others were more equivocal on the issue, and suggest that vaccines should not be considered for children until more data on long-term outcomes was available

> “I’ve read a lot of stuff lately about, you know … the long-term effects on people saying there’s not enough data. Why would you get a vaccine to a child, because you know we know how reacts in adults, what if there are adverse effects in children and like how could that work?” (Participant 21, Male, 20s)

In this sense, hesitancy in regard to children’s vaccines was similar to the hesitancy encountered in regard to adult vaccinations, which we have discussed elsewhere as ‘vaccine delay’ - that is where people are postponing the decision pending further information about the safety and efficacy of the vaccine.^15^ Vaccine delay was common to many participants:

> “There is not enough data to show how effective the vaccines are for children or what the implications may be and so maybe waiting for more government information and scientific data to backup that it’s important that children get vaccinated – before we make these decisions.”

Participants also discussed how long they felt they may need in order to decide whether vaccination for COVID-19 or children was something that should be pursued. Some felt it was dependent on when there was sufficient evidence on the safety of the vaccines in children or in general, although they tended to be unspecific as to what that evidence would consist of. Some suggested that they preferred to wait up to a year to see what the data showed or until then and “take it from there”:

> “This vaccine **-** which again has not been fully trialed on humans - I’d be very, very reluctant to [have my children take it]. … I have got a 6-year-old and a 13-year old … for at least a year and see the outcomes, if they can actually report any study outcomes.” (Participant 9, Male, 40s)

> “Those having [*sic*] children are thinking probably like myself, leave it at the moment, let’s see how we go 6 months to 12 months see in the data what’s happening and take it from there. It’s not fully tested at the moment” (Participant 6, Male, 40s)

These concerns tended to emphasise that the vaccines, in their view, had not been “fully tested” yet. As one parent put it: “I don’t want my son to be part of a giant experiment” (Participant 15, Female, 40+)

### Local social norms as a driver of hesitancy

Social norms, particularly local social norms – defined here as the views and beliefs of immediate family, friends and other social contacts – appeared to strongly influence people’s views. This included those who had children in the family:

> “Speaking to friends with children we seem to all feel similar. We wouldn’t want our children to be vaccinated, because we feel that if they get Covid hopefully they won’t be too ill.” (Participant 15, Female, 40+)

> “I have a young nephew, who spends a lot of time with my elderly parents… and the consensus in our family is that no he shouldn’t have it, and the consensus amongst friends who have children is also hesitancy to do this [vaccinate children] … Its far too early and unnecessary and the focus should be on the distribution and efficacy of vaccines amongst adults, and the booster campaign” (Participant 13, Male, 40s)

As the above quotes suggest, for a number of participants, speaking to influential, close others generated familial norms or friendship group norms, mostly around hesitancy or opposition to vaccination in children, including for some of the reasons discussed previously – such as uncertainty around COVID-19 in children (“hopefully they won’t be too ill”) and lower risk tolerance for unknown longer term effects (“it’s far too early”).

Other participants suggested that, in their view, the decision to make vaccines available to children would be met with wider opposition (including amongst parents), thus implying the absence of a more general social norm in favour of childhood COVID-19 vaccination:

> “I think a lot of people would be upset if they started saying you know that our children are going to have their nasal flu jab and we’re going to be offering a Covid jab as well in schools” (Participant 3, Male, 20s)

> “It [a vaccine] shouldn’t be promoted to children, I think is the government were to roll it out a lot of parents would have [*sic*] objection against that.” (Participant 2, Male, 30s)

### Association of the vaccine program with government’s handling of the pandemic

As with our previous research on attitudes to COVID-19 vaccines in adults, the extent of participants hesitancy related to the extent to which they saw the vaccines as being associated with the UK government.^15^ That is, those who were particularly hesitant about, or even opposed to, vaccination in children, tended to frame the vaccination program, including its potential extension to include children, in relation to, or as part of, the broader political context.

As discussed above, some participants were strongly opposed to COVID-19 vaccines. These participants were more vocal about their distrust in government, and the role of government in the potential extension of the use of vaccines in children. To return to a quote above (see: *COVID-19 vaccination in children as a “minefield”*), some participants were opposed to what they saw as a premature, and potentially unnecessary roll-out or “encroachment” “by stealth” something which one participant defined as “sinister” (Participant 13, Male, 40s).

Other participants who were hesitant but not strongly opposed to the idea, also framed their views in relation to what they saw as past government failures on COVID-19 policies, including England’s contact tracing system (‘Track and Trace’):

> “How can you trust the government or how much confidence do the public have with the government, now that the damage has been done, how can the public restore confidence … it was due to the failure of Dido Harding taking the contact tracing where did that go wrong?.. And now what we’ve noticed with the government, are parents prepared to take a risk for their own children?” (Participant 2, Male, 30s)

> “If they [the government] were having to vaccinate children, they were planning to do some kind of rollout of testing in schools, but they couldn’t even organise that. Like it was literally left up to schools … I think it does come back down to that all of the systems that are in place are really shoddy and like test and trace we know, has been proven it doesn’t work they spent billions [of pounds] on it … it ultimately comes down to trust (Participant 3, Male, 20s)

### Vaccinating children as a way to protect society (as collective responsibility)

A number of participants argued that vaccinating children might be a way to contribute to the overall population (‘herd’) immunity, and thus prove to be of benefit in society. In our previous research, on attitudes to COVID-19 vaccines in adults, we found a general distinction between those who framed vaccination as a collective act and those who framed vaccination as an individual act, with the latter tending to be more hesitant towards vaccines than the former.^15^ Amongst the data reported here, we also found that vaccination in children also tended to be framed as either an individual act – something that was the choice of the individual child’s parent(s) (see below) – or as a *collective act* – that is, as a way to protect others. Amongst our sample, few parents tended to discuss the wider, population benefits. The one exception was a parent – as discussed above (see: *COVID-19 vaccination in children as a “minefield”*), who acknowledged that childhood vaccination might “help get us out of the pandemic” but who also felt that “it’s much harder when it’s *your* children (Participant 15, Female, 30s)

Others in the group, non-parents, framed vaccination as way of protecting transmission to the more vulnerable in society, including their grandparents, thereby implying that they felt that although children may not “suffer” from COVID-19, they can spread it nonetheless:

> “I think it would be a good idea to vaccinate children. I know they say children don’t suffer so much when they get the virus if they catch it, but then to me its who they interact with at the end of the day, so you know they are going to go home to their parents who then go to work for example, or they are going to see their grandparents – and so to me I would be better if it was rolled out to try and flatten it down as much as possible.” (Participant 4, Male, 40s).

However, these participants often caveated or prefaced their views by emphasising that they themselves were not parents, and as such stated or implied that they had less say (or stake) in the decision-another important theme.

### Children’s vaccination as parental choice (as individual responsibility)

Another theme that emerged from the discussion was that parents were seen to be particularly important stakeholders. In our previous research on attitudes to COVID-19 vaccinations for adults,^15^ those who were more hesitant about the vaccines tended to frame their decision of whether or not to get a vaccine more as an individual choice. Conversely, those who tended to be more accepting (less prone to vaccine delay) tended to frame vaccination as a collective choice, corresponding with attitudes towards other COVID-19 measures like contact tracing compliance.^12^ Similarly, we found that hesitancy around whether or not children should be vaccinated was often framed in terms of vaccination as an individual choice – in this instance the choice of the individual parents. Those without children, often suggested that they were not “in a position to comment”:

> “I don’t have any kids and so I am really not in a position to comment or judge. I think it is really up to the parents on this one. You just have to put your trust in the science on this one. It’s a tricky one, if I did have kids, I would have to give it some serious consideration.

> “I can’t really comment on this either since I’m not a parent, it’s better to leave this decision to those who have children I think” (Participant 19, Male, 20s)

> “I don’t have children, so it’s easy for me to say if I did [what I would do]. I would have to know what benefits there were for the very young” (Participant 14, Female, 30s)

Other participants acknowledged that the decision was high-stakes, and carried a significant burden of responsibility (see also: *COVID-19 vaccination in children as a “minefield”*):

> “It is quite concerning when it’s your children you are responsible for their health and want the best for them – and you don’t want to make the wrong decision for them.” (Participant 5, Female, 20s)

One distinction that some participants made, was between the ability of older children to be able to make informed decisions for themselves, compared to younger children who were too young to understand the issue:

> “It comes down to people’s perception of like, you know, they are children and they can’t make decisions and the parents have to make decisions on whether they want to or not, whereas teenagers actually can form their own decision (Participant 3, Male, 20s)

## DISCUSSION

### Main findings of this study

This study found that participants framed COVID-19 vaccination in children as a complex issue, or “minefield”. Although a spectrum of views was found, from those largely in favour to those largely opposed to the idea of offering vaccines for children, most participants tended to be uncertain or hesitant about the idea, concluding that there was no straightforward answer. All parents in the group expressed considerable hesitancy, concluding that they would unlikely decide to have their child(ren) vaccinated were it to be made available at this point in time. The main reason cited was that they felt more information or evidence on the safety and efficacy of the vaccines in children was needed before they could decide.

Six main themes, or factors, shaping public attitudes to COVID-19 vaccines were identified. Firstly, there was uncertainty over whether children can catch, transmit or be severely harmed by COVID-19. This uncertainty partly reflected genuine scientific uncertainty that still exists, particularly around children’s role in transmission,^17^ but also may have been compounded by the confusion caused by changing messages and policies they experienced (e.g. around school testing and isolation policies). In the face of such uncertainty, participants tended to couch their views in affective terms (of a ‘feeling’ they had). Secondly, there was generally a lower risk tolerance for unknown longer-term effects of the vaccine in children. Whereas participants generally felt children were less susceptible to COVID-19, they felt they were more susceptible to long-term potential side effects of the vaccine, compared to adults. Parents suggested that they needed to see more evidence of testing and safety in children in order to feel confident. Thirdly, local social norms were a driver of hesitancy. Participants were strongly influenced by their own social networks, including for parents, other parents, where for many there is currently a culture of hesitancy around vaccination for COVID-19 in children. Fourthly, participants views were often framed in terms of trust in government; specifically, the extent to which they felt that the UK government could be trusted to successfully extend the vaccination programme to children (based on what they perceived as past failures over, for example, contact tracing). Fifthly, those who were more in favour, tended to emphasise the potential roll of COVID-19 vaccines for children in reducing overall infection rates, possibly by bringing up population (‘herd’) immunity. In this sense, individual vaccinations were framed as a collective act.^15^

Conversely, many participants also framed children’s vaccination as one of individual choice and responsibility. Non-parents tended to emphasise that the overall issue of whether vaccination should be made available for children was one that parents had a greater say or stake in. Parents tended to emphasise how difficult the issue was and how much responsibility they felt over the potential decision of whether or not they would have their child vaccinated. Thus, many may have a lower risk tolerance meaning that even those parents very accepting of vaccination in adults were more undecided or hesitant over whether vaccination in children was currently desirable.

### What is already known on this topic

Recent polls suggest that a majority of the public, including amongst parents, are in favour of offering COVID-19s to children are largely in favour of vaccine, these on their own fail to reveal the underlying complexity of the issue (for example via the use of binary response categories). ^8 9 10^. Survey research is also beginning to reveal some of the main reasons behind public attitudes to COVID-19 vaccination in children, with the most common reasons in favour being to prevent the spread of COVID-19 and to prevent children from catching COVID-19 and the main reasons against the idea being concerns over long-term effects of the vaccine on their child’s health, concerns over side-effects, a lack of knowledge of whether the vaccines had been tested in children.^10^

### What this study adds

This qualitative study is one of the first to document in-depth the reasons behind people’s attitudes towards the issue of COVID-19 vaccines in children, and, drawing on the UK context, suggests that for many people there is considerable uncertainty, complexity and ultimately hesitancy around the issue. Follow up research will be conducted as part of this longitudinal study to explore the extent to which attitudes towards COVID-19 vaccinations in adults evolves over time in much the same way as did attitudes towards COVID-19 vaccinations in adults, whereby a social norm around acceptance grew significantly, as vaccinations became increasingly approved for use in adults and as uptake increased in a growing number of countries globally.

### Limitations of this study

Strengths and limitations of the overall study have been discussed in previous publications.^11 12^ As with all qualitative studies, the generalizability of the findings is limited. As such, the strength of the study lies in its ability to reveal the complexity of the reasoning behind the extent to which people are supportive of COVID-19 vaccines in children, rather than in deriving proportions or gauging public sentiment overall. Additionally, due to the rapid nature of the call for participation from the participant pool, the sample size was slightly smaller than in previous rounds of data collection – although the total sample was deemed sufficient for the purposes of the analysis.

## Conclusions

Participants views on COVID-19 vaccinations in children reveal a complex issue, with many people, and particularly parents, tending to be quite uncertain or even hesitant about the issue. However, COVID-19 attitudes are rapidly changing, and so while currently no social norm around vaccination in children exists, as has been seen with attitudes towards vaccinations in adults,^15^ after they are approved for use in more countries including the UK, and as more under-18s start to get vaccinated, a more pro-vaccination norm may emerge. From a public health policy perspective, if and when COVID-19 vaccines are made readily available in the UK and other countries where they are not already available.

## Data Availability

Ethical restrictions related to participant confidentiality prohibit the authors from making the data set publicly available. During the consent process, participants were explicitly guaranteed that the data would only be seen my members of the study team. For any discussions about the data set please contact the corresponding author, Simon Williams.

## References

1. World Health Organization (2021) https://covid19.who.int/ https://covid19.who.int/ (accessed 27 July 2021)

2. UK Government (2021) GOV.UK Coronavirus (COVID-19) in the UK, vaccinations in the UK. https://coronavirus.data.gov.uk/details/vaccinations (accessed 27 July 2021)

3. Aschwanden, C. (2021) Five reasons why COVID herd immunity is probably impossible. Nature (News feature) https://www.nature.com/articles/d41586-021-00728-2. (accessed 21 July 2021).

4. Plotkin S, Levy P (2021) Considering Mandatory Vaccination of Children for COVID-19 Pediatrics, 147 (6) e2021050531; DOI: https://doi.org/10.1542/peds.2021-050531

5. Velavan, T. P., Pollard, A. J., & Kremsner, P. G. (2020). Herd immunity and vaccination of children for COVID-19. International journal of infectious diseases : IJID : official publication of the International Society for Infectious Diseases, 98, 14–15. https://doi.org/10.1016/j.ijid.2020.06.065

6. Gostin LO, Salmon DA, Larson HJ. Mandating COVID-19 Vaccines. JAMA. 2021;325(6):532–533. doi:10.1001/jama.2020.26553.

7. Opel DJ, Diekema DS, Ross LF. Should We Mandate a COVID-19 Vaccine for Children? JAMA Pediatr. 2021;175(2):125–126. doi:10.1001/jamapediatrics.2020.3019

8. Nolsoe E (2021) By 53% to 18%, parents with underage children say they would get them vaccinated against COVID-19. YouGov. https://yougov.co.uk/topics/health/articles-reports/2021/07/05/53-18-parents-underage-children-say-they-would-get (accessed 21 July 2021).

9. Office of National Statistics (ONS) (2021) COVID-19 Schools Infection Survey: England, Round 5. May 2021. https://www.ons.gov.uk/peoplepopulationandcommunity/healthandsocialcare/conditionsanddiseases/bulletins/covid19schoolsinfectionsurveyengland/round5englandmay2021 (accessed 21 July 2021).

10. IPSOS MORI (2021) 3 in 4 Britons support offering children the vaccine. https://www.ipsos.com/ipsos-mori/en-uk/3-4-britons-support-offering-children-vaccine (accessed 27 July 2021)

11. Williams S. N, Armitage C. J, Tampe T, Dienes K. (2020) Public perceptions and experiences of social distancing and social isolation during the COVID-19 pandemic: a UK-based focus group study. BMJ Open;10:e039334.

12. Williams, SN, Armitage, CJ, Tampe, T, Dienes K (2020) Public attitudes towards COVID-19 contact tracing apps: A UK-based focus group study. Health Expectations 24, 337–385 https://doi.org/10.1111/hex.13179

13. Williams S. N, Armitage C. J, Tampe T, Dienes K. (2020) Public perceptions of non-adherence to COVID-19 measures by self and others in the United Kingdom. medRxiv 2020.11.17.20233486; doi: https://doi.org/10.1101/2020.11.17.20233486

14. Gale, N. K., Heath, G., Cameron, E., Rashid, S., & Redwood, S. (2013). Using the framework method for the analysis of qualitative data in multi-disciplinary health research. BMC Medical Research Methodology, 13(1), 1–8. https://doi.org/10.1186/1471-2288-13-117

15. Williams S. N, Dienes K (2020) Public attitudes to COVID-19 vaccines: A qualitative study,medRxiv 2021.05.17.21257092; doi:https://doi.org/10.1101/2021.05.17.21257092

16. Slovic, P., Finucane, M.L., Peters, E. and MacGregor, D.G. (2004), Risk as Analysis and Risk as Feelings: Some Thoughts about Affect, Reason, Risk, and Rationality. Risk Analysis, 24: 311–322. https://doi.org/10.1111/j.0272-4332.2004.00433.x

17. Fischer, A. (2020) Resistance of children to Covid-19. How?. Mucosal Immunol 13, 563–565. https://doi.org/10.1038/s41385-020-0303-9

